# A Physiologically-Based Pharmacokinetic Model for Tuberculosis Drug Disposition at Extrapulmonary Sites

**DOI:** 10.1101/2022.12.31.22284083

**Authors:** Aparna Ramachandran, Chetan J. Gadgil

## Abstract

Tuberculosis (TB) is a leading cause of mortality due to an infectious agent. TB primarily targets the lungs but in about 16% cases can affect other organs as well, giving rise to extrapulmonary TB (EPTB). However, an optimal regimen for EPTB treatment is not defined. While the recommended treatment for most forms of EPTB is the same as pulmonary TB, pharmacokinetics of EPTB therapy are not as well-studied. To address this gap, we formulate a whole-body physiologically-based pharmacokinetic (PBPK) model for EPTB that for the first time includes the ability to simulate drug concentrations in the pleura and lymph node. Using this model, we estimate the time-dependent concentrations, at potential EPTB infection sites, of the 4 first-line anti-TB drugs: Rifampicin, Ethambutol, Isoniazid and Pyrazinamide. We utilise reported plasma concentration kinetics data to estimate model parameters for each drug, and validate our model using reported concentration data not used for model formulation or parameter estimation. Model predictions match the validation data and reported PK parameters (*Cmax, tmax*) for the drugs. The model also predicts Ethambutol, Isoniazid and Pyrazinamide concentrations in the pleura that match reported experimental values from an independent study. For each drug, the model is used to simulate the time-dependent drug concentrations at various EPTB sites. Predicted drug concentrations are compared to their critical concentration. Simulations suggest that while Rifampicin and Isoniazid concentrations are greater than critical concentration values at most EPTB sites, the concentrations of Ethambutol and Pyrazinamide are lower than their critical concentrations at most EPTB sites.

## 1. Introduction

Tuberculosis (TB) is an infectious disease caused by the pathogen *Mycobacterium tuberculosis*. It is spread when aerosol droplets containing the bacteria are inhaled by a susceptible individual^1^. The World Health Organization (WHO) predicts that in 2019, around 10 million people fell ill with TB and 1.4 million people died from it^2^. This large mortality number has only continued to increase with an estimated 1.6 million deaths in 2021^3^. According to the WHO, TB is one of the major causes of mortality among adults due to a single infectious agent, only surpassed by COVID-19^3^. Progress in reducing global TB burden has been adversely affected due to the COVID-19 pandemic. There has been a significant reduction in the number of newly-diagnosed cases, as well as in access to TB diagnosis and treatment^3^. In addition, it has been observed that heavy use of corticosteroids in COVID-19 patients suppressed the immune system and led to the reactivation of latent TB^4^.

Pulmonary TB is the most common presentation of the disease^5^. The standard treatment of drug-sensitive TB consists of an regimen with four first-line drugs: rifampicin, isoniazid, pyrazinamide, and ethambutol^6^. TB therapy is composed of an intensive phase of two months with isoniazid, rifampicin, pyrazinamide, and ethambutol, followed by a continuation phase of at least four months with isoniazid and rifampicin^6^. However, resistance to these antimicrobials can emerge, giving rise to drug-resistant TB.

In addition to pulmonary TB, the disease can affect organ systems such as the lymphatic system, the gastrointestinal system, the central nervous system and others, collectively referred to as extrapulmonary TB (EPTB). In 2019, about 16% of the reported 7.1 million cases were classified as EPTB^2^. Lymph node TB is the most frequent form of EPTB, followed by pleural TB. Other locations of EPTB manifestations include the genitourinary tract, skin, bones or joints, and meninges^5,7^.

In spite of accounting for almost one-sixth of the total TB notifications, EPTB continues to be overlooked. The recommended treatment for most forms of EPTB is the same as pulmonary TB^7^. EPTB therapy is not as well-studied as pulmonary TB^7^, and given its prevalence, further improvement and standardisation of EPTB treatment is necessary. Physiologically-based pharmacokinetic (PBPK) modelling can be employed to predict the *in vivo* distribution of anti-TB drugs. Such mathematical tools can complement preclinical studies of drugs, reducing both time and resources utilized in drug development.

Several whole-body PBPK models evaluating the pharmacokinetics of anti-TB drugs in adults have been developed. These have been summarized in Table S1. None of these models simulate the pharmacokinetics of all four first-line drugs. To our knowledge, only one study models EPTB treatment through a PBPK model. The model is used to predict the time-dependent tissue concentration of delamanid relative to its MIC at extrapulmonary sites such as the brain, heart and liver^8^. However, this model is a single drug distribution model that does not include the pleura and lymph nodes, which are important sites of EPTB.

We develop a PBPK model to study the temporal distribution of the four first-line anti-TB drugs in humans, specifically at extra-pulmonary sites. In our work we formulate a whole-body human PBPK model that for the first time (i) includes drug distribution to pleura and lymph node and (ii) simulates the time-dependent tissue concentrations of all four primary anti-TB drugs: rifampicin, ethambutol, isoniazid, pyrazinamide. We use the model simulations to examine the predicted concentrations at extrapulmonary sites relative to the effective concentrations of the drugs. We employ available concentration data of the drugs to calibrate and independently validate our model. Two distinctive features of our model are the analysis of drug pharmacokinetics in the pleural fluid and the lymph nodes, prevalent sites of EPTB. Slow and rapid acetylation of isoniazid are considered as discrete cases. Such PBPK studies can help evaluate the effectiveness of currently prescribed treatment regimens for EPTB and help develop and evaluate potential new protocols.

## 2. Materials and Methods

### 2.1 Model Formulation

The structure of the whole-body PBPK model is adapted from Lyons et al^9^. A mass balance on the drug in individual compartments leads to a system of ordinary differential equations (ODEs). The rate of change of amount of drug *A*_*T*_ in a tissue/organ T is described using the following general equation:

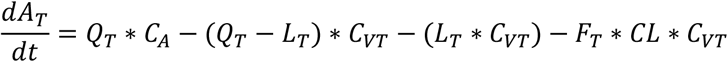

Here, *Q*_*T*_ (L/hr) is the flow rate to and from the tissue/organ, *L*_*T*_ (L/hr) is the lymph flow rate from tissue/organ, *C*_*A*_ (μg/mL) is the drug concentration in arterial blood, *CL* (L/hr) is total systemic clearance of the drug, *F*_*T*_ is the fraction of total clearance apportioned to T (set to zero if no clearance occurs in T), and *C*_*VT*_ (μg/mL) is the drug concentration exiting T with *C*_*VT*_ *= C*_*T*_*/P*_*T*_, where *P*_*T*_ is the tissue:blood partition co-efficient. Amount of drug in tissue T is *A*_*T*_ *= C*_*T*_*\*V*_*T*_, where *V*_*T*_ is the volume of T.

### 2.2 Model Parameters

#### 2.2.1 Physiological Parameters

Cardiac output and afferent lymph flow rate were based on previously reported values. Tissue volumes, blood flow rates and lymph flow rates were taken as fraction of body weight, cardiac output and afferent lymph flow rate respectively, for a 70 kg man. These fractional values were also taken from previous reports. All reported values are summarized in the Supplementary Tables S2 and S3. No lymph flow was considered from spleen and bone compartments. Fractional tissue volume, blood flow rate and lymph flow rate for the compartment representing the ‘rest of the body’ were calculated as the remaining fraction.

#### 2.2.2 Tissue:Plasma Partition Coefficients

To determine the tissue/plasma partition coefficients (*P*_*T*_) for the four drugs, the Rodgers and Rowland method^10,11^ was employed. Reported rat tissue composition data for the tissues and organs was used. Values for different properties of blood cells and plasma were assumed to be those used by Rodgers and Rowland^10,11^. Values for various chemical and biological properties of the drugs are listed in Supplementary Table S4. The median of all *P*_*T*_ values for a drug was taken as the *P*_*T*_ for the rest of the body. The calculated partition coefficient values are listed in Supplementary Table S5.

#### 2.2.3 Estimation of Other Parameters

By fitting our model output to available time-dependent plasma concentration and lung concentration data, pharmacokinetic (PK) parameters and lung tissue/blood partition coefficient (*P*_*Lu*_) values were estimated. Absorption rate (*k*_*a*_) and systemic clearance (*CL*) of the drug were the PK parameters estimated using plasma concentration-time data. Model output plasma concentrations for oral doses of rifampicin (450 mg), ethambutol (400 mg), isoniazid (300 mg) and pyrazinamide (2000 mg) were fitted to reported plasma concentration data^12–16^ for each drug. In case of plasma, since several data sets were available, separate sets of studies were used for model calibration and model validation. Lung tissue concentration values at different time points for rifampicin, isoniazid and pyrazinamide were obtained from a study by Prideaux et al^17^. Given the limited number of studies reporting lung tissue concentration, this data for each of these drugs was split into two sets: one to estimate the lung tissue/blood partition coefficient (*P*_*Lu*_) values, and another to validate this calculated parameter value. The *P*_*Lu*_ values were estimated for rifampicin and pyrazinamide and were used for all further simulations. The *P*_*Lu*_ value for isoniazid and ethambutol were computed using the Rodgers and Rowland method, using the same methodology as for other tissues and organs. Fractional renal clearance values were taken from previous reports, as listed in the SI. Gut reabsorption rate for rifampicin was set to 0.17 hr^-1^ based on the reported value^9^.

The optimization function used was weighted sum of squares error, defined in the Supplementary Information (SI). Goodness-of-fit plots were constructed to quantify the match of model simulations to clinical data, including data that was not used for parameter estimation. Estimated parameters were used unchanged in all further simulations.

### 2.3 Simulation

The system of ODEs was numerically solved using *ode23s* in MATLAB 2020b to obtain time-series drug concentration prediction for the 4 first-line TB drugs in different tissues and organs. *fitnlm* was used for parameter estimation. The predictions of the model were validated using available drug concentration data. Simulated plasma concentrations for standard oral doses of rifampicin (600 mg), ethambutol (1200 mg), isoniazid (300 mg) and pyrazinamide (1500 mg) were plotted along with reported plasma concentration data^12,13,18–27^ for each drug dose respectively. At the start of the dosing regimen, there is a transient lasting several days when the average daily concentration increases and then reaches a steady state. While simulating drug concentrations in EPTB organs, unless mentioned otherwise, 24-hour time profiles of the drug concentration at day 7 after start of the drug treatment were used to represent the long-term daily profile of drug concentration. In the pleura, day 8 concentrations were simulated since reported data for rifampicin, isoniazid and pyrazinamide was for the 8^th^ day after the start of the dosing regimen. Simulated pleural drug concentrations for standard oral doses of rifampicin (600 mg), ethambutol (1200 mg), isoniazid (300 mg) and pyrazinamide (1500 mg) were plotted along with reported concentration data^28–30^ for each drug dose. None of the data used for model calibration were used for validation of the simulations. Plots comparing simulated and experimental values were constructed wherever *in vivo* data was available.

Model output concentrations were compared to the WHO-prescribed critical concentration for each drug^31,32^, which is the lowest drug concentration that inhibits growth of 95% of M. tuberculosis strains isolated from untreated patients, but does not inhibit growth of strains isolated from patients resistant to that drug^32^. As pyrazinamide activity depends on the surrounding pH, in addition to the critical concentration at pH 6.0, MIC at pH 5.5-5.7^33^ was also considered for comparison.

## 3. Results

### 3.1 PBPK model incorporating EPTB sites

We extended a previously reported^9^ PBPK model to include two major sites of EPTB, viz, pleural space and lymph nodes. This new model is shown in Figure 1. We introduce a ‘lymph node’ compartment similar to the venous and arterial blood compartments, to represent the lymphatic system. This compartment collects afferent lymph from organs (except bone and spleen) and drains into the venous blood compartment. The pleural space is represented as a sub-compartment of the lung that receives filtrate from the lungs and drains to the lymph node. This representation of physiology is consistent with reports that pleural fluid is a microvascular filtrate flowing in through the parietal pleural capillaries^34,35^ and removal of this fluid from the pleural space occurs mainly via lymphatic stomata in the parietal pleura^35,36^.

**Figure 1:** Schematic diagram of PBPK model incorporating pleural and lymph node compartments

The model consists of 17 perfusion-limited, well-stirred tissue compartments connected by blood and lymph flow, and parameterised by known physiological values from literature as well as calculated tissue:plasma partition coefficients and pharmacokinetic parameters. The model is described using a system of 18 ordinary differential equations (ODEs), listed, along with the parameters used, in the Supplementary Text. Concentration-time profiles in various tissues and organs were simulated for the four first-line anti-TB drugs: Rifampicin, Ethambutol, Isoniazid and Pyrazinamide. Additionally, as the pharmacokinetics of Isoniazid vary significantly depending on the acetylator status of the individual, we model two cases: slow (SA) and fast acetylators (FA).

Oral administration of drug was assumed, represented by introduction of the drug into the gut compartment with first-order absorption. First-order elimination was assumed. Rifampicin is known to undergo enterohepatic circulation^37^. This was modelled as first order re-absorption of the drug from the gut lumen.

### 3.2 Model Calibration and Validation

#### 3.2.1 Model Calibration

The model output plasma concentration and lung concentration of the drugs were fit to reported data. The drug absorption rate, systemic clearance and lung tissue/blood partition coefficient (*P*_*Lu*_) values were estimated by minimizing the weighted sum of squared error between model prediction and reported drug concentrations. Figure 2 shows the fitted curves and reported data for plasma concentration. Goodness-of-fit plots for predicted (from model) and observed (from literature) drug concentrations in the plasma are shown in Figure S1. Fitted lung concentration curves and goodness-of-fit plots comparing observed and predicted concentrations can be found in Figure S2. Fast acetylators have a higher *CL* value than slow acetylators, as expected. The experimental data used here for model calibration was not used further for validation.

**Figure 2:** Model output plasma concentrations for oral doses of rifampicin (450 mg), ethambutol (400 mg), isoniazid (300 mg) and pyrazinamide (2000 mg) were fitted to reported plasma concentration data for each to predict parameters. Reported data used is listed in Materials and Methods.

#### 3.2.2 Model Validation

The developed model was validated using reported plasma concentration data for each drug to assess the efficacy of the formulated model. Data different from that used for parameter estimation was used for validation. The obtained simulation results are shown in Figure 3. Goodness-of-fit plots for predicted and observed drug plasma concentrations can be found in Figure S3. Parameters are unchanged from calibrated values. The only parameter that is varied is the dose administered. It is seen that the model is able to simulate data not used in model calibration, indicating the robustness of model prediction.

**Figure 3:** Simulated plasma concentrations for standard oral doses of rifampicin (600 mg), ethambutol (1200 mg), isoniazid (300 mg) and pyrazinamide (1500 mg) plotted along with reported plasma concentration data for each drug dose. Reported data used is listed in Materials and Methods. The model parameters are listed in Table S1 in the SI. Data used for calibration was not used for model validation.

### 3.3 Model Prediction of Drug Concentration

#### 3.3.1 Pleura

Lymph node TB and pleural TB are the most common forms of EPTB. To address pleural TB, we incorporated a pleural fluid compartment in the model to predict the concentration of first-line anti-TB drugs and hence approximate their effectiveness against pleural TB. The parameters values are not changed from the calibrated values. Dose administered is set to the experimentally reported value. The simulated day 8 drug concentrations in the pleura are shown in Figure 4. For drugs other than Rifampicin, the simulations and reported data seem to be in good agreement. The model over-predicts the pleural concentration of rifampicin. This is consistent with the observed trend of rifampicin exhibiting lesser penetration into the pleura than the other first-line drugs^38^. Predicted concentrations of pyrazinamide and ethambutol are below their prescribed critical concentrations, while those of Rifampicin and Isoniazid are well above their critical concentrations.

**Figure 4:** Simulated pleural drug concentrations for standard oral doses of rifampicin (600 mg), ethambutol (1200 mg), isoniazid (300 mg) and pyrazinamide (1500 mg) plotted along with reported concentration data for each drug dose respectively. The model parameters are listed in Table S1 in the SI. The grey dashed line represents the critical concentration of each drug (also MIC at pH 5.5-5.7 for PYZ). We use day 7 simulation for all drugs except ethambutol for which we simulate day 2 concentration as data reported by Elliott et al. are for day 2.

#### 3.3.2 Lung Tissue

We use the model to simulate lung tissue concentrations and compared to experimental data. The parameters values used in the simulation are not changed from the calibrated values. Dose is set to experimentally reported value. Lung tissue concentration-time data for rifampicin, isoniazid and pyrazinamide for output validation was taken a study by Prideaux et al^17^. The simulated lung tissue concentrations are shown in Figure 5. The agreement between the model output and the reported data is reasonable given that the experimental data was not used for model calibration. The model predicts that while Rifampicin and Isoniazid concentrations are above the critical concentration for a large percentage of time, the concentration of Ethambutol is below the critical concentration for a large fraction of time, and the concentration of Pyrazinamide is always below its critical concentration.

**Figure 5:** Simulated lung tissue drug concentrations for standard oral doses of rifampicin (600 mg), ethambutol (1200 mg), isoniazid (300 mg) and pyrazinamide (1500 mg) plotted along with reported concentration data for each drug dose. The grey dashed line represents the critical concentration of each drug (also MIC at pH 5.5-5.7 for PYZ). Data used for parameter estimation is not used here.

#### 3.3.3 Extrapulmonary Sites

Time-dependent concentrations of the four drugs were simulated at other potential sites of EPTB and compared to their critical concentrations to predict their effectiveness at combating infection at these sites. Figure 6 shows the day 7 concentrations of the drugs, following the initial treatment. For reference, the critical concentrations are also shown. Drug concentrations at different EPTB sites relative to that in the lung compartment over time are shown in Figure S4.

**Figure 6:** Simulated day 7 drug concentrations at various sites of EPTB for standard oral doses of rifampicin (600 mg), ethambutol (1200 mg), isoniazid (300 mg) and pyrazinamide (1500 mg) compared to the critical concentration of each drug (also MIC at pH 5.5-5.7 for PYZ). The same parameter values from model calibration are used here.

While isoniazid achieves a peak concentration that is quite high relative to its critical concentration in both fast and slow acetylators, rapid metabolisers are unable to maintain this as the drug is rapidly eliminated. Rifampicin appears to achieve a peak concentration higher than its critical concentration in all cases, and stays above this concentration for a large fraction of the time in all organs. According to our predictions, ethambutol is unable to reach a concentration higher than the critical concentration in brain, kidney, bone and skin compartments. An important point to consider for ethambutol and rifampicin is their limited penetration into the cerebrospinal fluid (CSF), especially when meningeal inflammation is not too prominent^39^. As this is not taken into account, the model probably over-predicts the in vivo concentrations of these drugs in the brain and the actual concentrations are even smaller. In a study where patients were administered 600 mg of rifampicin via infusion approximately 3 hours before surgery, the highest observed concentration in normal brain tissue was just 0.56 μg/mL^40^. While IV administration results in higher concentrations than those attained by oral administration of the same dose, this value was much less than that predicted by our model around this time point. As they are moderately lipophilic small molecules, pyrazinamide and isoniazid do not face this issue of poor CNS penetration and hence are vital in the treatment of TB meningitis^41^. Pyrazinamide, in its current dose, fails to attain its critical concentration in any of the organs in our simulations. A study on 7H10 agar determined pyrazinamide MIC to be around 18-22 μg/mL at a pH of 5.5^42^. The estimated concentrations of pyrazinamide are lower than even this lowered MIC in all tissues.

## 4. Discussion

The objective of our work was the development of a whole-body PBPK model to study the disposition of the four first-line anti-TB drugs rifampicin, ethambutol, isoniazid, and pyrazinamide in an EPTB context, focusing on lymph node TB and pleural TB, the predominant forms of EPTB. We added two compartments to a previously reported whole body-PBPK model: a consolidated lymph node compartment and pleura. Following the estimation of tissue:plasma partition coefficients and PK parameters, our model was able to estimate the in vivo concentration-time profiles of each drug in different tissues/organs. Validation of the projected concentrations would require tissue concentration data that is unavailable for most cases due to the need for invasive sampling. Hence, we use plasma concentrations and limited lung tissue concentration data for this purpose.

As we assume drug susceptible TB in our model, we consider critical concentrations of the drugs (as prescribed by WHO) as these concentrations inhibit 95% (90% for pyrazinamide) of wild strains of our target bacteria. We also assume that there are no significant drug-drug interactions between the four drugs and that the pharmacokinetics of a drug are independent of the concentration of the others. It has been shown that Rifampicin can increase levels of cytochromes P450, increasing its own metabolism^43^. We do not incorporate the autoinduction demonstrated by rifampicin. Additionally, we consider the pharmacokinetics of the drugs to be similar in people with and without TB and do not consider age-dependent PK properties.

Apart from these assumptions, another limitation of our model is assumption of a consolidated single lymph node compartment. There are around 600 lymph nodes in the body and a combined compartment is clearly not an accurate representation of this anatomy. Moreover, drug distribution to all lymph nodes will not be equal. Nonetheless, studying the drug disposition to the lymph nodes is pertinent when discussing EPTB treatment. As such, our model may be regarded as the first step in the incorporation of lymph nodes in whole-body PBPK models for TB.

Despite these assumptions, our model, calibrated using one set of experimental data, shows good agreement with data from a different source. It is also able to predict Ethambutol, Isoniazid and Pyrazinamide concentrations in the pleura that match reported experimental values from a study that was not used for either calibration or validation. The expected maximum plasma concentration (*Cmax*) of the drugs administered at their standard adult doses are 8-24 μg/mL for rifampicin, 2-6 μg/mL for ethambutol, 3-5 μg/mL for isoniazid and 20-50 μg/mL for pyrazinamide achieved at expected times (*tmax*) of 2 hours, 2-3 hours, 0.75-2 hours, and 1-2 hours respectively^44^. Our simulations are consistent with these observations.

We used our model to predict time-dependent concentrations of the four first-line anti-TB drugs in extrapulmonary organs. Rifampicin attained a peak above its critical concentration in all simulated cases. However, in the pleural fluid compartment, our model seems to greatly over-predict the time-dependent concentrations of the drug. Reported pleural fluid drug concentrations seem to barely cross the critical concentration threshold. In a study, the authors observed that rifampicin levels in the pericardial effusion of pericardial tuberculosis patients were lower than expected. They hypothesize that a plausible explanation for this is the thickening of pericardial layers, or fibrosis^45^. Something equivalent could be a reason for the dramatically lower concentrations in the pleura as pleural fibrosis is a known complication of tuberculous pleural effusion (TPE)^46^. Additionally, as it is highly protein-bound, rifampicin exhibits limited penetration into CSF and treatment of tuberculous meningitis might benefit from an increased dosage^47^. In vivo, rifampicin bioavailability is also affected by its metabolism auto-induction.

Ethambutol fails to reach a concentration above its prescribed critical concentration in the brain, kidney, bone and skin compartments. It has been observed that CSF ethambutol concentrations are too low to be quantifiable in healthy volunteers, and is detectable but in low amounts in TBM patients with inflamed meninges^39^. Hence, with rehabilitation, the contribution of ethambutol to therapy becomes negligible. In the other tissues/organs its maximum concentration exceeded the critical concentration.

Isoniazid concentrations are above the critical threshold in all compartments, in both rapid and slow acetylators. However, the *Cmax* and time above the critical concentration vary greatly. As expected, slow metabolisers attain a greater peak concentration *Cmax* in all compartments and are able to maintain the drug concentration above the effective concentration for a relatively longer period of time. Therefore, customization of dosage of isoniazid based on the acetylator status of the individual may prove to be beneficial. Isoniazid has been reported to be antagonistic towards rifampicin^48,49^ and pyrazinamide^48,50^.

In our simulations, pyrazinamide does not achieve a maximal concentration higher than its MIC or critical concentration in any compartment. Environmental pH has been seen to play an important role in the sterilizing activity of pyrazinamide. Hence, we take pH into account while setting effective concentration cut-off for this drug. Pyrazinamide’s anti-bacterial activity has been shown to increase with decreasing pH values^33^. A more detailed analysis of the relation of pyrazinamide’s sterilising activity to the environmental pH and how this affects treatment is provided in the Supplementary text.

In many organs, we observe that the simulated *Cmax* of the drug does not cross the critical concentration barrier, or the drug concentration does not stay above this threshold for a very long period of time. However, this cocktail of drugs is universally prescribed for all forms of TB. Synergistic action of the drugs that enhances the anti-mycobacterial effects of these drugs is not accounted for in the model. Additionally, in the host, the bacteria experience immune-mediated stress, resulting in greater in vivo drug effect against the pathogen. Hence, a quantitative system pharmacology (QSP) model including the PK/PD of the drug as well as the immune processes in the body might provide a more holistic picture.

PBPK modelling offers a number of advantages. Inter-individual variability within a demographic, or across different demographics can be integrated into a model. This technique can be immensely useful in pre-clinical drug development. It can also be valuable in evaluating potential new regimens, since it is desirable that drugs reach their targeted concentrations in particular organs while avoiding problems associated with higher doses. A more detailed and intensive in vivo analysis of drug concentrations and PK at EPTB sites is required to understand the effectiveness of the currently prescribed regimen, and can help optimize dosing study of EPTB. The model and results presented here will serve as a starting point for such studies of drug distribution at EPTB sites.

## Study Highlights

### What is the current knowledge on the topic?

Several whole-body PBPK models evaluating the pharmacokinetics of anti-TB drugs in adults have been developed. However, these models do not address the two major EPTB sites: pleura and lymph nodes, and are focus on pulmonary TB.

### What question did this study address?

This study seeks to formulate a whole-body PBPK model to predict drug concentrations at various EPTB sites. We use this new model to predict pharmacokinetics of drugs in extrapulmonary organs.

### What does this study add to our knowledge?

To our knowledge, this is the first model to study pharmacokinetics for pleural and lymph node TB. It provides reasonable predictions in some cases, such as concentrations of most of the drugs in the pleura. Data is required to validate predictions at other sites.

### How might this change clinical pharmacology or translational science?

EPTB treatment needs to be addressed and optimized. PBPK modelling can be used to design new, more effective, treatment regimens. This model will serve as the starting point for development of models for EPTB treatment.

## Data Availability

All Matlab codes used to produce the data in the present study are available upon reasonable request to the authors

## Acknowledgements

C.G. acknowledges funding from CSIR-NCL and DBT (BT/PR40128/BTIS/37/43/2022). A.R. acknowledges funding from CSIR through a research fellowship. C.G. and A.R. thank Rajesh Gokhale for useful discussions on EPTB; and the organisers of the India-EMBO symposium on Mycobacterial heterogeneity and host tissue tropism.

## Author Contributions

C.G. designed the research. A.R performed the research. A.R. and C.G. analysed the data and wrote the manuscript.

## Supplementary Information Titles

1. PBPK Model Equations
2. Objective Function Used for Minimization to Estimate Parameters
3. Supplementary Tables Table S1: A summary of relevant whole-body PBPK models for adults incorporating anti-TB drugs Table S2: Physiological parameters for the assumed male individual Table S3: Tissue-wise physiological parameters values Table S4: Chemical and biological properties of the 4 first-line anti-TB drugs Table S5: Tissue-wise calculated partition coefficient values Table S6: Weights assigned to different experimental data points during model calibration Table S7: Predicted PK parameters for each drug
4. Supplementary Figures Figure S1: Model Calibration – Goodness-of-fit plots for predicted and reported plasma concentrations for oral doses of rifampicin (450 mg), ethambutol (400 mg), isoniazid (300 mg) and pyrazinamide (2000 mg). Concentration vs time predictions are shown in Figure 2. Figure S2: Model Calibration – Curve-fitting predicted lung tissue concentrations rifampicin and pyrazinamide to reported concentrations alongside goodness-of-fit plots for predicted and observed lung tissue concentrations. Oral doses of 600 mg rifampicin and 1500 mg pyrazinamide were administered. Figure S3: Model Validation – Goodness-of-fit plots for predicted and observed drug plasma concentrations in simulations. Oral doses of rifampicin (600 mg), ethambutol (1200 mg), isoniazid (300 mg) and pyrazinamide (1500 mg) were simulated. Concentration vs time predictions are shown in Figure 3. Figure S4: Ratio of time-dependent drug concentrations at different EPTB sites to that in the lung compartment over time. The dashed line indicates a ratio of 1.

## Notes

**Conflict of interest:** All authors declared no competing interests for this work

### Competing Interest Statement

The authors have declared no competing interest.

### Funding Statement

This study was funded by Council of Scientific and Industrial Research in the form of a fellowship to Aparna Ramachandran; and Department of Biotechnology in the form of a research grant to Chetan Gadgil (BT/PR40128/BTIS/37/43/2022)

