## Supplementary Information for "A Physiologically-Based Pharmacokinetic Model for Tuberculosis Drug Disposition at Extrapulmonary Sites"

#### 1. PBPK Model Equations

**Venous blood:**

$$V_V \frac{dC_V}{dt} = \sum_T (Q_T * C_{VT}) + (L_{LN} * C_{VLN}) - Q_C * C_V$$

**Arterial blood:**

$$V_A \frac{dC_A}{dt} = (Q_C - L_{Lu}) * C_{VLu} - (Q_C - L_{Lu}) * C_A$$

**Lungs:**

$$V_{Lu} \frac{dC_{Lu}}{dt} = Q_C * C_V - (Q_C - L_{Lu}) * C_{VLu} - (L_{Lu} - Q_{Pl}) * C_{VLu} - Q_{Pl} * C_{VLu}$$

**Pleura:**

$$V_{Pl} \frac{dC_{Pl}}{dt} = Q_{Pl} * C_{VLu} - Q_{Pl} * C_{Pl}$$

**Non-eliminating tissues/organs with afferent lymph (Brain, Heart, Adipose, Muscle, Skin, Others):**

$$V_T \frac{dC_T}{dt} = Q_T * C_A - (Q_T - L_T) * C_{VT} - L_T * C_{VT}$$

**Non-eliminating tissues/organs without afferent lymph (Bone, Spleen):**

$$V_T \frac{dC_T}{dt} = Q_T * C_A - Q_T * C_{VT}$$

**Kidney:**

$$V_{Kd} \frac{dC_{Kd}}{dt} = Q_{Kd} * C_A - (Q_{Kd} - L_{Kd}) * C_{VKd} - L_{Kd} * C_{VKd} - f_R * CL * C_A$$

**Gut:**

$$V_{Gu} \frac{dC_{Gu}}{dt} = Q_{Gu} * C_A - (Q_{Gu} - L_{Gu}) * C_{VGu} - L_{Gu} * C_{VGu} + k_a * A_D + k_r * A_{GL}$$

**Liver:**

$$V_{Li} \frac{dC_{Li}}{dt} = Q_{LA} * C_A + (Q_{Sp} - L_{Sp}) * C_{VSp} + (Q_G - L_G) * C_{VG} - (Q_{Li} - L_{Li}) * C_{VLi} - L_{Gu} * C_{VGu} - (1 - f_R) * CL * \frac{Q_{LA} * C_A + Q_{Sp} * C_{Sp} + Q_G * C_G}{Q_{Li}}$$

**Gut Lumen (GL):**

$$\frac{dA_{GL}}{dt} = (1 - f_R) * CL * \frac{Q_{LA} * C_A + Q_{Sp} * C_{Sp} + Q_G * C_G}{Q_{Li}} - k_r * A_{GL} - k_F * A_{GL}$$

**Lymph Node:**

$$V_{LN} \frac{dC_{LN}}{dt} = \sum_T (L_T * C_{VT}) - L_{LN} * C_{VLN}$$

Here, $Q_T$ (L/hr) is the flow rate to and from a tissue/organ "T", $L_T$ (L/hr) is the lymph flow rate from tissue/organ "T", $C_A$ (µg/mL) is the drug concentration in arterial blood, $Q_{Pl}$ is the flow rate of the pleura, *CL* (L/hr) is total systemic clearance of the drug, $F_T$ is the fraction of total clearance apportioned to T (if any), and $C_{VT}$ (µg/mL) is the drug concentration exiting T with $C_{VT} = C_T/P_T$, where $P_T$ is the tissue:blood partition co-efficient for T. Amount of drug in tissue T is $A_T = C_T*V_T$, where $V_T$ is the volume of T.

#### 2. Objective Function Used for Minimization to Estimate Parameters

Least squares method is used by the *fitnlm* function during model calibration. Hence, the sum of squares of the offsets of experimental data points from the model simulated concentration curve is minimised, where we assign weight to certain data points. This function for *n* reported data points is:

$$\sum_{i=1}^{n} w_i (y_i - f(x_i))^2$$

$y_i$ is the measured value of the dependent variable, $f(x_i)$ is the model predicted value and $w_i$ is the weight assigned to $i^{th}$ observation. The values assigned for $w_i$ are listed in Table S6.

#### 3. Lymph Node Compartment

To our knowledge, the integration of a lymph node compartment is a novel addition to the study of tuberculosis using PBPK. Many studies have incorporated a lymph node compartment in their PBPK model to evaluate the pharmacokinetics of different substances such as peptides[1], monoclonal antibodies[2,3], nanoparticles[4,5] as well as small molecules[6]. A recent PBPK study based on non-human primates has demarcated the lymph node network into five major regions, which then drain into the thoracic duct[7].

#### 4. Relation of Pyrazinamide Activity to Environmental pH and its Effect on Treatment

Environmental pH has been seen to play an important role in the sterilizing activity of pyrazinamide. Pyrazinamide's anti-bacterial activity has been shown to increase with decreasing pH values[8]. Pyrazinamide is thought to target non- or slowly-reproducing bacteria in acidic compartments such as the macrophage phagosome[9,10]. While immature phagosomes have a pH of 6.2, post bacilli internalization by macrophage, acidification occurs, resulting in a phagosomal pH of pH 4.5 to 5.0[11]. However, this notion is contradicted by the finding that macrophage vesicles containing *M. Tuberculosis* bacteria were not acidic[12]. It has also been suggested that the drug exhibits antimicrobial activity against extracellular slow-replicating bacteria in the epithelial lining fluid[13]. Poor treatment response to pyrazinamide in animal infection models such as mice and guinea pigs, with neutral to alkaline lesion pH, provide further evidence in favour of the enhanced activity of the drug in acidic conditions[10,11]. This observed higher pH in TB lesions in guinea pig and murine models does not appear to be an impediment to treatment with pyrazinamide in humans though, as shown in a study by Kempker et al. where a majority of the lesion samples studied by them (8 out of 10 patients) had an acidic pH ($\leq 5.5$)[14]. As stated by Srivastava et al., pH in human TB cavities varies around 5.5, while that in murine TB models is higher[15]. This is a probable cause for differential outcomes in the two cases.

### 5. Supplementary Tables

**Table S1[†]:** A summary of relevant whole-body PBPK models for adults incorporating anti-TB drugs

| | Drug | Type of TB | Features of the Model | References |
|---|---|---|---|---|
| 1 | Isoniazid | Pulmonary | • Describes NAT2-dependent pharmacokinetics of isoniazid and its metabolites<br>• Includes acetylator status (fast, intermediate, slow)<br>• Includes PD | [17] |
| 2 | Isoniazid | Pulmonary | • Employs two coupled PBPK models: one for a lactating mother and one for her infant to study drug exposure in the infant from drug intake by the mother<br>• Includes acetylator status (fast and slow) | [18] |
| 3 | Isoniazid | Pulmonary | • Assessment of potential drug-drug interactions with CYP2C19 and CYP3A4 substrates<br>• Includes acetylator status (fast and slow) | [19] |
| 4 | Ethambutol | Pulmonary | • Considers scenarios that reflect different stages of PBPK model development to evaluate drug pharmacokinetics | [20] |
| 5 | Rifampicin | Pulmonary | • Recognizes and models differences in rifampicin pharmacokinetics after a single dose in healthy, TB and cirrhosis populations | [21] |
| 6 | Rifampicin, Ethambutol | Pulmonary | • Structured model with two organisms: lactating mother and nursing infant | [23] |

[†] In these models, human physiology is described by representing organs and tissues as compartments. The number of compartments varies, depending on the modelling approach adopted. Each compartment can be homogenous and well-stirred or consist of sub-compartments. A notable exception is the representation of the lung as a multi-compartment permeability-limited organ[16]. These PBPK studies simulate the time-dependent concentrations of a single drug[17–22], as well as multiple drugs[16,23–26], for first-line and many second- and third-line anti-TB drugs. The models that study first-line drugs do not include EPTB sites as their focus is pulmonary TB. To our knowledge, only one study models EPTB treatment through a PBPK model[22].

| | | | | |
|---|---|---|---|---|
| 7 | Rifampicin, Isoniazid, Pyrazinamide, Ethambutol | Pulmonary | • Properties predicted from mice were used to deduce parameters and predict lung:plasma ratio in humans which were compared to biopsy data from patients | [24] |
| 8 | Bedaquiline, Delamanid, Isoniazid, Rifapentine | Pulmonary | • Simulates the long-acting administration of select anti-TB drugs for LTBI treatment<br>• Includes acetylator status | [25] |
| 9 | Delamanid | Pulmonary and Extra-pulmonary (Brain, Heart, Liver) | • Simulated concentrations of the drug in the lung, brain, liver, and heart and found them to be higher than the estimated effective concentration | [22] |
| 10 | Rifampicin, Ethambutol, Isoniazid, Itraconazole, Erythromycin, Clarithromycin, Pyrazinamide | Pulmonary (Lungs) | • Incorporates a multi-compartment permeability-limited lung model instead of a single homogeneous lung compartment | [16] |
| 11 | Bedaquiline, Clofazimine, Cycloserine, Isoniazid Ethambutol, Ethionamide, Kanamycin, Pyrazinamide, Rifampicin, Linezolid | Pulmonary (Lungs) | • Model accuracy assessed using drug plasma concentrations and lung tissue concentrations | [26] |

**Table S2:** Physiological parameters for the assumed male individual

| Parameter | Value | References |
|---|---|---|
| Body weight | 70 kg | Assumption |
| Cardiac output | 5200 mL/min | [27] |
| Afferent lymph flow rate | 8 L/day | [28] |
| Gut lumen transit rate | 0.252 $hr^{-1}$ | [29] |

**Table S3:** Tissue-wise physiological parameters values

| Organ/Tissue | Symbol | Volume[2,27,30] (as fraction of Body Weight) | Blood Flow Rate[27,29] (as fraction of Cardiac Output) | Lymph Flow Rate[31] (as fraction of Afferent Lymph Flow) |
|---|---|---|---|---|
| Lungs | Lu | 0.0076 | - | 0.03 |
| Brain | Br | 0.02 | 0.12 | 0.0105 |
| Adipose | Ad | 0.2142[a] | 0.05 | 0.128 |
| Heart | Hr | 0.0047 | 0.04 | 0.01 |
| Muscle | Mu | 0.4 | 0.17 | 0.16 |
| Bone | Bo | 0.1429[a] | 0.05 | 0 |
| Skin | Sk | 0.0371 | 0.05 | 0.0703 |
| Kidney | Kd | 0.0044 | 0.19 | 0.085 |
| Spleen | Sp | 0.0026 | 77/5200[e] | 0 |
| Gut | Gu | 0.0171 | 1100/5200[e] | 0.12 |
| Liver | Li | 0.0257 | $Q_{LA} + Q_{Gu} + Q_{Sp}$ | 0.33 |
| Hepatic Artery | LA | - | 0.06 | - |
| Lymph Node | LN | 0.274/70[b] | - | - |
| Arterial Blood | A | 1.8/70[c] | - | - |
| Venous Blood | V | 3.6/70[c] | - | - |
| Others | Oth | 0.04264[d] | 0.04365[f] | 0.0562[g] |
| Pleura | $Q_{Pl}$ | 0.3 mL $kg^{-1}$*[32] | 0.15 mL $kg^{-1}$ $h^{-1}$*[32] | - |

a: Density = Mass/Volume. We assume that Density ≈ 1 $g/cm^3$ and so Mass ≈ Volume, except for adipose where density = 0.916 $g/cm^3$ and for bone where density = 1.92 $g/cm^3$

b: Taken from Shah & Betts, 2012 (Combined volume of LNs = 274 mL)

c: Taken from Igari et al. (Volume of arterial blood = 1.8 L, venous blood = 3.6 L)

d: Others = 1 – (Sum of other compartments) = 1 – 0.9576 = 0.0424

e: Taken from Davies & Morris, 1993

f: Others = 1 – (Sum of other compartments) = 1 – 0.95635 = 0.04365 (Pleura fraction is considered negligible)

g: Others = 1 – (Sum of other compartments) = 1 – 0.9438 = 0.0562

**Table S4:** Chemical and biological properties of the 4 first-line anti-TB drugs

| | Rifampicin | Ethambutol | Isoniazid | Pyrazinamide[a] |
|---|---|---|---|---|
| **Compound type** | Zwitterion[33] (group 1) | Diprotic base[34] | Monoprotic base[34] | Neutral[34] |
| **Acid dissociation constant (pKa)** | pKa1 = 1.7, pKa2 = 7.9[33] | pKa1 = 6.5, pKa2 = 9.55[34] | 1.82[34] | 0.5[35] |
| **logPo:w** | 2.7[33] | -0.3[36] | -0.7[37] | -0.6[35] |
| **logPvo:w** | 1.7 | -1.7 | -2.1 | -2 |
| **BP** | 0.9[38] | 0.99[#] | 1* | 1* |
| **$Kpu_{BC}$** | 5.19 | 1.30[39] | 1.33 | 1.11 |
| **$Ka_{BC}$** | 8.33 | 1.31 | – | – |
| **fu** | 0.15[40] | 0.75[34] | 0.95[34] | 0.9[34] |
| **fR** | 0.07[41] | 0.79[41] | Fast: 0.07[42]<br>Slow: 0.29[42] | 0.09[41] |

a: Pyrazinamide is hydrophilic nature[43]. Lipoproteins usually binds with hydrophobic drugs[44]. Hence, it is assumed that pyrazinamide interacts majorly with albumin instead of lipoproteins.

logPo:w – n-octanol:water logP
logPvo:w – vegetable oil:water logP is required to estimate adipose tissue Kp and is calculated using the linear regression relationship proposed by Leo et al.[45] between logPo:w and logPvo:w as experimental values were not found.. The equation used here is an adaptation of this relationship, by Poulin and Theil[46], *logPvo:w = 1.1115 * logPo:w − 1.35*
BP – Blood:plasma ratio of the drug
KpuBC – Blood cell:plasma water unbound drug concentration ratio. Is calculated[47] as *(H − 1 + BP)/(fu*H)* where haematocrit H is taken to be 0.45[46], except for ethambutol for which experimental value is available
$Ka_{BC}$ – Ka is the association constant of basic/zwitterionic drugs with acidic phospholipids of a tissue. $Ka_{BC}$ corresponds to Ka for blood cells and is calculated using $Kpu_{BC}$[48]. Ka values for drugs are not available readily and hence are approximated as KaBC.
fu – fraction of drug unbound in the plasma
fR – fractional renal clearance
*: BP value for isoniazid and pyrazinamide were assumed to be 1. In $P_T$ calculation, the B:P (blood/plasma partition coefficient) for isoniazid and pyrazinamide have been set to 1 as this experimental data is unavailable in literature. This is based on the reported assumption that for drugs that are distributed homogenously into tissues, B:P can be taken to be 1[46].
#: BP value was calculated from $Kpu_{BC}$

**Table S5:** Tissue-wise calculated partition coefficient values

| Organ/Tissue | Rifampicin | Ethambutol | Isoniazid | Pyrazinamide |
|---|---|---|---|---|
| Lungs | 0.9341[a] | 4.3579 | 0.7662 | 1.3379[a] |
| Brain | 0.2290 | 1.8014 | 0.7537 | 0.7184 |
| Adipose | 0.1890 | 0.4586 | 0.1543 | 0.1503 |
| Heart | 1.0187 | 3.0400 | 0.7550 | 0.7243 |
| Muscle | 0.6968 | 2.6942 | 0.7208 | 0.6868 |
| Bone | 0.3166 | 1.3699 | 0.4330 | 0.4163 |
| Skin | 0.6282 | 1.9808 | 0.6674 | 0.6497 |
| Kidney | 2.1789 | 5.2877 | 0.7441 | 0.7127 |
| Spleen | 1.3990 | 3.9689 | 0.7605 | 0.7260 |
| Gut | 1.0812 | 3.1812 | 0.7429 | 0.7140 |
| Liver | 1.9704 | 5.0521 | 0.7212 | 0.6887 |
| Lymph Node | 1.2116 | 3.6471 | 0.7556 | 0.7210 |
| Others[b] | 1.0812 | 3.1812 | 0.7441 | 0.7140 |

a: Calculated by model fitting

b: Median of all other values

**Table S6:** Weights assigned to different experimental data points during model calibration

**Rifampicin**

Fig. 2 – Acocella, 1978

| 1 | 2 | 3 | 4 | 5 | 6 | 7 | 8 |
|---|---|---|---|---|---|---|---|
| 0.0238 | 0.0238 | 0.2381 | 0.0238 | 0.0238 | 0.0238 | 0.0238 | 0.0238 |

Fig. 2 – Furesz, 1970

| 1 | 2 | 3 | 4 | 5 | 6 |
|---|---|---|---|---|---|
| 0.4763 | 0.0238 | 0.0238 | 0.0238 | 0.0238 | 0.0238 |

Fig. S3 – Prideaux et al., 2016

| 1 | 2 | 3 | 4 | 5 | 6 | 7 |
|---|---|---|---|---|---|---|
| 0.6667 | 0.0333 | 0.0333 | 0.0333 | 0.0333 | 0.0333 | 0.1668 |

**Ethambutol**

Fig. 2 – Strauch et al., 2011 (Etb-91-400B)

| 1 | 2 | 3 | 4 | 5 | 6 | 7 | 8 | 9 | 10 | 11 |
|---|---|---|---|---|---|---|---|---|---|---|
| 0.0208 | 0.0208 | 0.0208 | 0.0208 | 0.3128 | 0.0208 | 0.0208 | 0.0208 | 0.0208 | 0.2083 | 0.0208 |

Fig. 2 – Strauch et al., 2011 (Etb-ref-400)

| 1 | 2 | 3 | 4 | 5 | 6 | 7 | 8 | 9 | 10 |
|---|---|---|---|---|---|---|---|---|---|
| 0.0208 | 0.0208 | 0.0208 | 0.0208 | 0.0208 | 0.1045 | 0.0208 | 0.0208 | 0.0208 | 0.0208 |

**Isoniazid (FA)**

Fig. 2 – Gallicano et al., 1994

| 1 | 2 | 3 | 4 | 5 | 6 | 7 | 8 | 9 | 10 | 11 | 12 |
|---|---|---|---|---|---|---|---|---|---|---|---|
| 0.0400 | 0.4000 | 0.0400 | 0.0400 | 0.0400 | 0.0400 | 0.0400 | 0.0400 | 0.0400 | 0.0400 | 0.0400 | 0.2000 |

**Isoniazid (SA)**

Fig. 2 – Gallicano et al., 1994

| 1 | 2 | 3 | 4 | 5 | 6 | 7 | 8 | 9 | 10 | 11 | 12 | 13 | 14 |
|---|---|---|---|---|---|---|---|---|---|---|---|---|---|
| 0.0357 | 0.5359 | 0.0357 | 0.0357 | 0.0357 | 0.0357 | 0.0357 | 0.0357 | 0.0357 | 0.0357 | 0.0357 | 0.0357 | 0.0357 | 0.0357 |

**Table S7:** Predicted PK parameters for each drug

| **Drug** | | **Absorption rate, $k_a$ ($h^{-1}$)** | **Systemic Clearance, $CL$ ($L\ h^{-1}$)** | **Lung tissue : Plasma Partition Coefficient, $P_{Lu}$** |
|---|---|---|---|---|
| Rifampicin | | 0.9095 | 9.1130 | 0.9336 |
| Ethambutol | | 0.2221 | 50.4888 | – |
| Isoniazid | Fast | 2.9895 | 24.5171 | – |
| | Slow | 4.1112 | 9.1799 | |
| Pyrazinamide | | 1.5148 | 4.3085 | 1.3798 |

#### 6. Supplementary Figures

**Figure S1:** Model Calibration – Goodness-of-fit plots for predicted and reported plasma concentrations for oral doses of rifampicin (450 mg), ethambutol (400 mg), isoniazid (300 mg) and pyrazinamide (2000 mg). Concentration vs time predictions are shown in Figure 2.

**Figure S2:** Model Calibration – Curve-fitting predicted lung tissue concentrations rifampicin and pyrazinamide to reported concentrations alongside goodness-of-fit plots for predicted[49] and observed lung tissue concentrations. Oral doses of 600 mg rifampicin and 1500 mg pyrazinamide were administered.

**Figure S3:** Model Validation – Goodness-of-fit plots for predicted and observed drug plasma concentrations in simulations. Oral doses of rifampicin (600 mg), ethambutol (1200 mg), isoniazid (300 mg) and pyrazinamide (1500 mg) were simulated. Concentration vs time predictions are shown in Figure 3.

**Figure S4:** Ratio of time-dependent drug concentrations at different EPTB sites to that in the lung compartment over time. The dashed line indicates a ratio of 1.
